# Oral Ketamine for the Treatment of Depression: A randomized controlled trial and meta-analysis

**DOI:** 10.1101/2025.03.21.25324252

**Authors:** Leo R. Silberbauer, Benjamin Eggerstorfer, Paul Michenthaler, Sabine Reichel, Thomas Stimpfl, Thomas Vanicek, Angela Naderi-Heiden, Siegfried Kasper, Rupert Lanzenberger, Gregor Gryglewski

**Author notes:** **Correspondence:** Gregor Gryglewski, MD, PhD, Department of Psychiatry and Psychotherapy, Medical University of Vienna, Austria, Waehringer Guertel 18-20, 1090 Vienna, Austria.

## Abstract

Ketamine represents a significant advancement in antidepressant therapy, but the commonly used intravenous and intranasal application routes currently limit its availability beyond specialized centers. By contrast, oral ketamine treatment might constitute an alternative that is widely available, easy to administer, and has potential advantages with regard to tolerability. Forty-five patients diagnosed with a moderate to severe depressive episode received either 1mg/kg peroral ketamine or 0.03mg/kg midazolam solution as an active comparator, administered six times over two weeks in a double-blind trial. While statistical significance for the primary endpoint of MADRS score reduction after one week was not achieved, response rates favored ketamine with a number needed to treat (NNT) of 4.6 (95%-CI: [2.4, 62.6] at this timepoint. Treatment was well-tolerated, with no serious adverse events reported, potentially due to the lower exposure to ketamine compared to its metabolite norketamine measured in plasma. In a total of 592 patients, meta-analysis of randomized controlled trials on oral ketamine treatment demonstrated its antidepressant efficacy with a NNT = 4.89 (95%-CI: [3.41, 8.66]) for response and a NNT = 9.10 (95%-CI: [5.43, 28.03]) for remission. Despite that the current trial did not meet its primary endpoint, the cumulative evidence up-to-date suggests that oral ketamine treatment leads to relevant improvements in the outcomes of patients with depression. In light of the ease of administration and high tolerability with oral application, this evidence may contribute to removing some of the obstacles that currently restrict the availability of antidepressant treatment with ketamine to specialized centers.

## INTRODUCTION

Alleviating the burden of depression remains a major challenge in psychiatry. While there is a wide array of evidence-based treatment options (Malhi and Mann, 2018), the majority of patients does not respond sufficiently to first-line antidepressant treatment (Bartova et al., 2019). Ketamine, with its rapid onset of efficacy in difficult to treat cases, constitutes a major advancement in antidepressant therapy as mounting evidence continues to substantiate its efficacy in uni- and bipolar depression (Fava et al., 2018; Phillips et al., 2019; Saelens et al., 2024; Zarate et al., 2012b). After the initial proofs of concept (Berman et al., 2000; Zarate et al., 2006), most trials have continued to study the effects of ketamine administered intravenously. The approval of intranasal esketamine by the U.S Food and Drug Administration (FDA) and the European Medicines Agency (EMA) for treatment resistant depression (TRD) (McIntyre et al., 2021) in 2019 has greatly extended its availability to patients in need. While the intranasal route facilitates drug application, the extensive monitoring of patients required due to side effects such as elevated blood pressure, sedation and dissociative states continues to limit its use to specialized centers (Kasper et al., 2021; Ross and Soeteman, 2020).

Peroral ketamine administration has proven to be effective in the management of chronic pain across several conditions and self-administration in the home setting was shown to be feasible (Peltoniemi et al., 2016). After ingestion, ketamine undergoes extensive first-pass metabolism which results in higher levels of metabolites (Fanta et al., 2015) that were previously implicated in its antidepressant effects (Yang et al., 2019) and might lack adverse effects and abuse potential (Zanos et al., 2016). Moreover, slow kinetics after oral application may be associated with increased tolerability (Bowdle et al., 1998). The ease of oral ketamine administration may further provide access to treatment for selected patient populations that are not eligible for intravenous or intranasal esketamine treatment, such as patients with uncontrolled arterial hypertension (Silberbauer et al., 2020) and patients in palliative care settings (Irwin et al., 2013).

To date, several case series and open label studies support the antidepressant effects of oral ketamine treatment (Meshkat et al., 2023). Moreover, oral ketamine was shown to have beneficial effects in the treatment of patients with chronic suicidality (Can et al., 2021; Dutton et al., 2022). Emerging evidence from randomized, controlled trials is promising, demonstrating the effectiveness of oral ketamine in TRD (Domany et al., 2019; Kheirabadi et al., 2020), in accelerating antidepressant response in combination with sertraline (Arabzadeh et al., 2018), and in patients with major depression, including those with chronic pain (Jafarinia et al., 2016) and those experiencing suicidal ideation (Seraj et al., 2025).

The accessibility and low cost of oral ketamine treatment warrants further investigation to substantiate its efficacy and safety across different patient populations and settings. In this randomized, controlled trial, we assessed the rapid efficacy, tolerability and pharmakokinetics of oral ketamine in patients diagnosed with major depressive or bipolar disorder suffering from a moderate to severe depressive episode. A meta-analysis of available randomized controlled trials was performed to contextualize our findings within the broader body of evidence.

## METHODS

### Participants

Patients were recruited from the in- and outpatient clinics of the Department of Psychiatry and Psychotherapy at the Medical University of Vienna. Patients diagnosed with a moderate to severe depressive episode in the scope of a major depressive disorder (MDD) or bipolar disorder were eligible for enrollment. Clinical diagnoses were supported by structured clinical interviews for DSM-IV (SCID-I). Inclusion criteria comprised a 17-item Hamilton Rating Scale for Depression (HDRS) score ≥19 and stable psychopharmacological treatment for at least 10 days. Inclusion did not require prior antidepressant treatment or evidence of treatment resistance. History of psychotic symptoms or substance abuse (except nicotine) were exclusion criteria. All patients provided written informed consent. All procedures were reviewed and approved by the ethics committee of the Medical University of Vienna (EK No. 1536/2016) and carried out according to the Declaration of Helsinki. This study was registered before the start of recruitment at clinicaltrials.gov (NCT02992496). Subjects were recruited between 24.04.2017 and 07.04.2021.

### Study design and medication

This trial followed a double-blind, randomized, controlled study design. Participants were randomly assigned to receive either 1mg/kg ketamine (ketamine hydrochloride, Hameln Pharma Plus GmbH) or 0.03mg/kg midazolam (Roche Pharma AG). An active control condition was chosen to optimize blinding and to mitigate the influence of unspecific effects on treatment outcome. Midazolam displays similar pharmacokinetic properties as ketamine and, thus, mimics the time course of its non-specific behavioral effects (Murrough et al., 2013). The study medication was administered six times over a period of 2 weeks with one or two days between each dose.

### Assessment of antidepressant efficacy

Clinician rated depression severity and antidepressant efficacy was assessed using the Montgomery-Åsberg Depression Rating Scale (MADRS) and the HDRS. Ratings were performed at baseline (screening), 24 hours after the first dosing and immediately before each subsequent treatment session. Follow-up assessments were performed one and two weeks after the last administration of the study medication. Efficacy ratings were performed by clinicians who were not involved in the administration of the study medication and the assessment of tolerability.

### Assessment of tolerability

Drug effects were assessed using the Clinician Administered Dissociative States Scale (CADSS) (Bremner et al., 1998) and the Altered States of Consciousness Rating Scale (5D-ASC, 11-ASC) 30 minutes after the first dosing (Studerus et al., 2010). Moreover, visual analog scales (VAS) were performed 30 minutes after the first and last administration to detect the emergence of most common adverse effects as reported by Murrough et al. (aan het Rot et al., 2010).

### Statistical analysis

All statistical analyses were performed using R (v4.0.2, https://www.R-project.org/). Descriptive statistics of demographic and clinical variables were compared between treatment groups. Reduction in MADRS total score one week after start of treatment was the primary endpoint of this study. An intention-to-treat analysis comprising all patients who received at least one dose was performed. A mixed effects model for repeated measures (baseline, 24 hours, 7 days and 11 days after first dose) was calculated using maximum likelihood estimation. The intercept was allowed to vary randomly. Treatment group, time of visit and their interaction were included as fixed effects and subject as random effect.

Secondary end points included the comparison of response (≥50% MADRS reduction compared to screening) and remission rates (MADRS <10) between treatment groups. Response and remission rates were each analyzed using the last observation carried forward (LOCF) method to account for missing data. Moreover, safety and tolerability after the first dose as assessed using the CADSS and the ASC were compared between treatment groups using independent two-sided t-tests. Values greater than 1 on the VAS were counted as adverse effects and frequencies between treatment groups after single and repeated dose were compared using Fisheŕs exact test. Exploratory correlational analyses were conducted to examine the association between subjective drug effects and antidepressant treatment response, and to explore the relationship between plasma levels, treatment efficacy, and tolerability (see supplement).

### Meta-analysis of RCTs assessing antidepressant efficacy of oral ketamine

The bibliographic databases PubMed, Scopus, PsycInfo, Web of Science and Google Scholar were systematically searched for randomized controlled trials that assessed the antidepressant efficacy of oral ketamine treatment in depression. As the MADRS and the HDRS was utilized to assess antidepressant efficacy across included studies, standardized mean difference (Cohen’s d) of the change in depression score between ketamine and control conditions was calculated (Morris and DeShon, 2002). Furthermore, meta-analyses based on risk differences (absolute risk reduction) in response and remission rates between treatment groups were calculated and reported as number needed to treat (NNT) for each study. Multilevel random effects models were employed with random effects specified at both the study level and within study level to account for multiple treatment groups within studies. Variance components were estimated using restricted maximum likelihood and the models were used to obtain summary effect sizes.

## RESULTS

Forty-seven participants met the inclusion criteria. Two subjects were lost to follow-up before the first dose, four subjects dropped out during double-blind treatment. The primary endpoint was analyzed using intention to treat analysis in all patients who received one dose (n= 45). Treatment groups did not differ in terms of age, sex and baseline depression severity (see Table 1).

**Table 1.** Demographics and clinical characteristics of patients (mean ± sd). Groups did not differ in terms of age, sex and baseline depression severity. MDD, Major depressive disorder; BD, Bipolar disorder MADRS, Montgomery-Åsberg Depression Rating Scale; HDRS17, 17-item Hamilton Depression Rating Scale; BDI, Beck Depression Inventory. SSRI, Selective serotonin reuptake inhibitor; SNRI, Serotonin-norepinephrine reuptake inhibitor; SARI, Serotonin antagonist and reuptake inhibitor; NaSSa, Noradrenergic and specific serotonergic antidepressant; NDRI, Norepinephrine-dopamine reuptake inhibitor.

|  | **Ketamine**<br>**(n=23)** | **Midazolam**<br>**(n=22)** |
| --- | --- | --- |
| Age (y) | 38.3 ± 13.4 | 43.3 ± 13.3 |
| Sex (f/m) | 11/12 | 9/13 |
| Diagnosis (MDD/BD) | 17/6 | 18/4 |
| MADRS | 31.9±3.4 | 31.7±7.3 |
| HDRS<sub>17</sub> | 24.1±3.4 | 23.7±5.0 |
| BDI | 29.7±6.9 | 29.5±8.9 |
| Current psychopharmacological treatment |  |  |
| None | 3 (13.0%) | 5 (22.7%) |
| SSRI | 8 (34.8%) | 7 (31.8%) |
| SNRI | 8 (34.8%) | 5 (22.7%) |
| SARI | 3 (13.0%) | 3 (13.6%) |
| NaSSa | 4 (17.4%) | 5 (22.7%) |
| NDRI | 6 (26.1%) | 5 (22.7%) |
| Tricyclic Antidepressant | 2 (8.7%) | 0 |
| Tetracyclic Antidepressant | 1 (4.3%) | 0 |
| Mood stabilizer | 8 (34.8%) | 5 (22.7%) |
| Second generation antipsychotic | 8 (34.8%) | 7 (31.8%) |
| Antipsychotic | 1 (4.3%) | 2 (9.1%) |
| Benzodiazepine | 5 (21.7%) | 3 (13.6%) |
| Benzodiazepine on demand | 5 (21.7%) | 3 (13.6%) |
| Antiepileptic drug | 5 (21.7%) | 2 (9.1%) |
| Stimulant | 2 (8.7%) | 1 (4.5%) |
| Other | 0 | 1 (4.5%) |

### Antidepressant efficacy: Primary endpoint

The linear mixed effects model showed significant main effects of time of visit (24 hours, 7 days, 11 days), but interaction effects (time of visit x treatment group) did not reach statistical significance at any time point after treatment initiation (p > 0.05, Figure 1). There were numerically higher reductions in depression rating scores in the ketamine compared to the midazolam treatment group.

**Figure 1.**
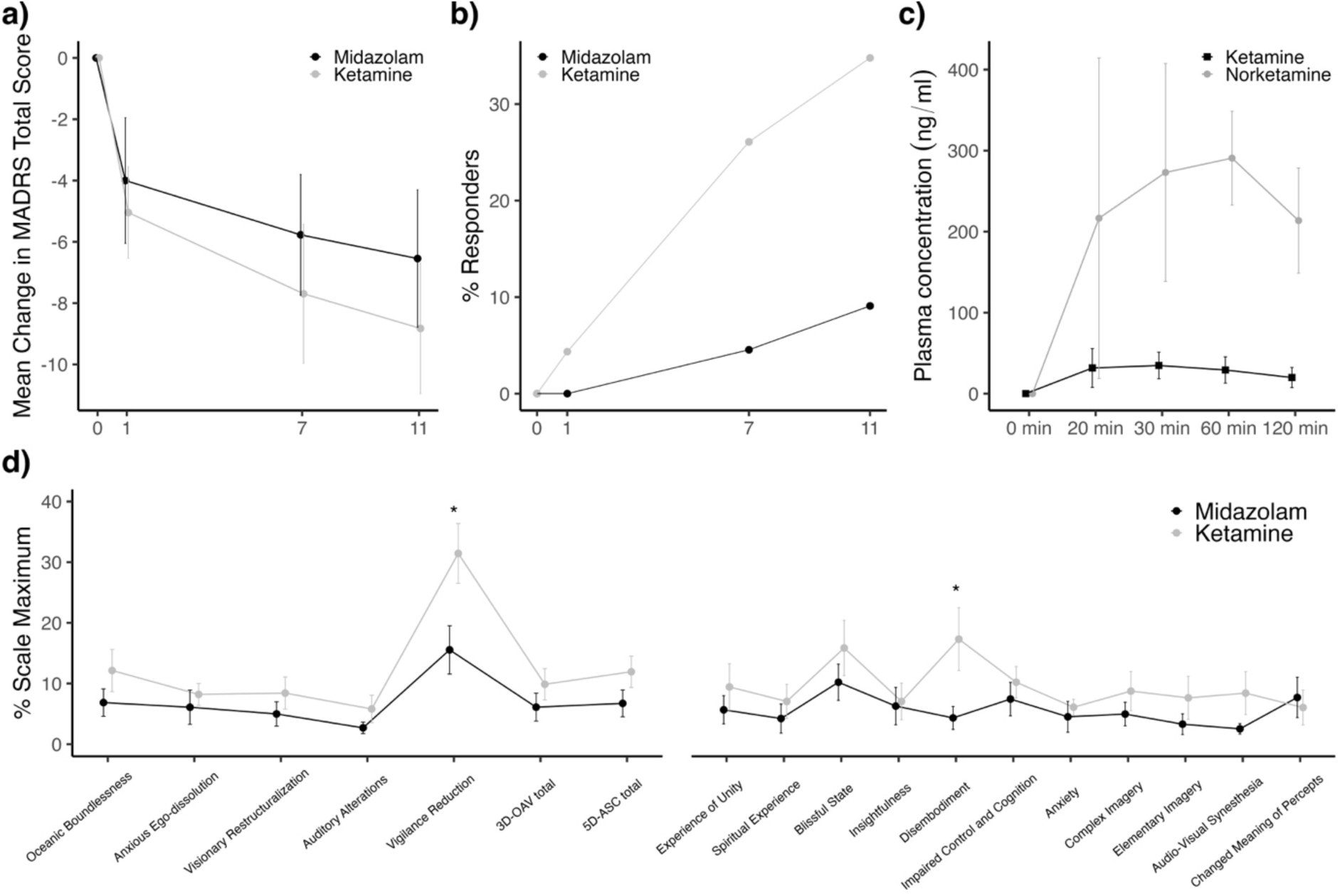
a) Mean change in MADRS total score from baseline at 24 hours (1 day), 7 days and 11 days after the first dose of repeated oral ketamine administration is displayed. We did not observe a statistically significant difference between ketamine and midazolam at any timepoint. Error bars indicate the standard error of the mean. b) The proportion of MADRS responders per group at each time point. c) Serial ketamine and metabolite plasma concentration. Median plasma concentration (ng/ml) is shown 20 to 120 minutes after the first dose. Error bars denote median absolute deviation. d) Drug effects on the Altered States of Consciousness Rating Scale (ASC) scale. Mean (± standard error of the mean) of the scalés maximum is displayed. Significant differences between ketamine and midazolam were found in “vigilance reduction” and “disembodiment”.

### Antidepressant efficacy: Secondary endpoints

A number needed to treat (NNT) for response of 4.6, 95% confidence interval (CI): [2.4, 62.6] after 7 days and 3.9, 95% CI: [2.1, 35.5] after 11 days was observed. There was no significant difference in response and remission rates between groups (see Table S1). Two weeks after completion of the double-blind treatment phase, we observed a response rate of 38.5% at the final visit among 39 patients, following naturalistic antidepressant treatment, which included open-label administration of peroral ketamine in some patients (mean ± sd MADRS score at final visit = 17.80 ± 10.47).

### Safety and tolerability

30 minutes after the first application, a higher Clinician Administered Dissociative States Scale (CADSS) total score was observed in the ketamine compared to the control group (t=-2.78, p<0.01) (see Table S2). In the ASC a higher vigilance reduction (t=-2.51, p=0.02) and experience of disembodiment (t=-2.36, p=0.03) was observed after ketamine (Figure 1).

Among the items assessed with the VAS 30 minutes after the first study drug administration, more patients in the ketamine group reported dizziness (p<0.01), poor coordination (p<0.05), poor concentration (p<0.01), and anxiety (p<0.05) after the first dose (Table 2). In contrast, the only side effect reported at a higher rate after the last dose of ketamine vs. midazolam was poor coordination (p<0.05).

**Table 2.**
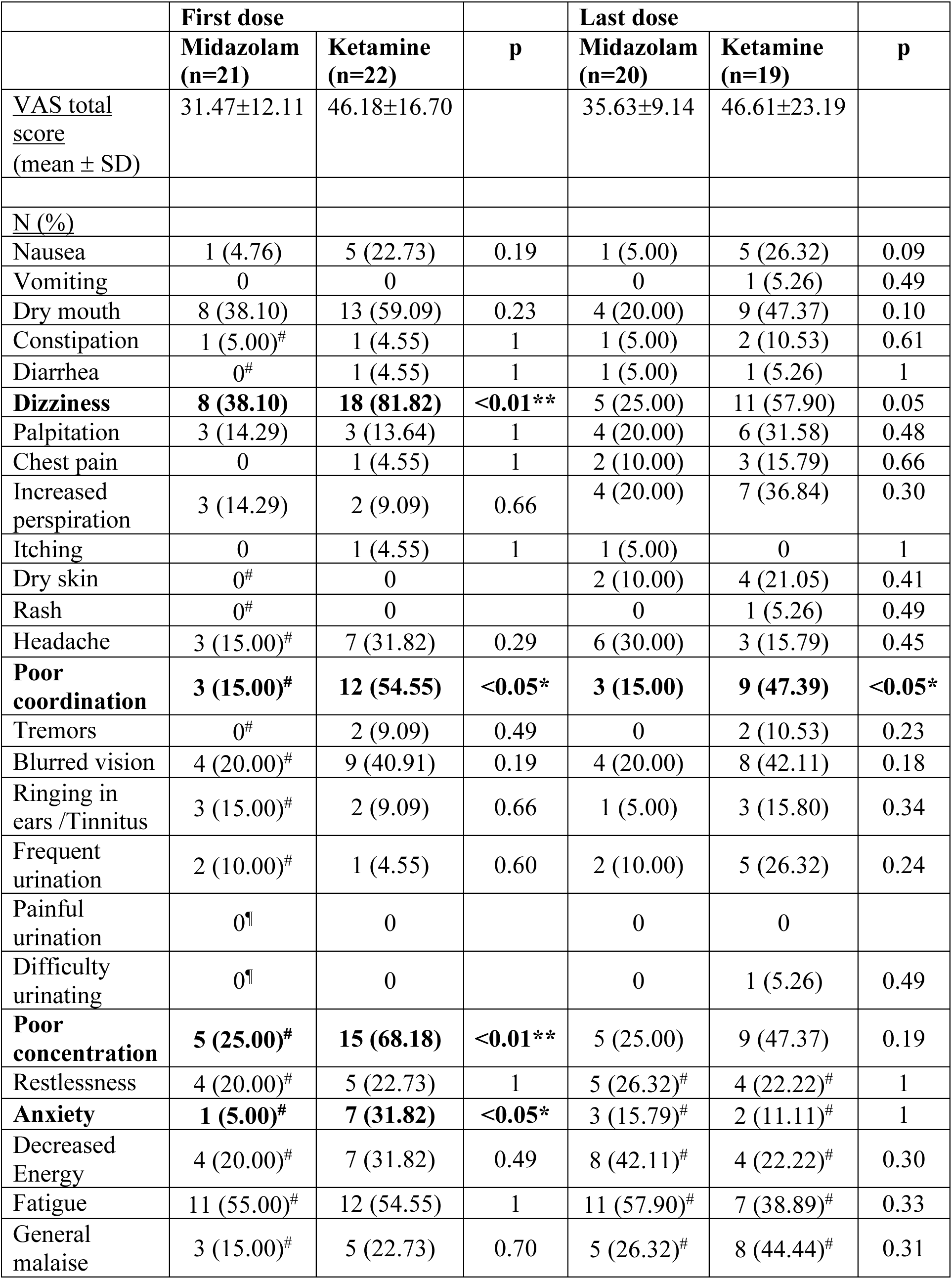
Results of the visual analogue scale (VAS) assessing commonly reported side effects as reported by Murrough et al. Counts of more than 1 on the VAS are reported as number of patients and percentages of the total number of patients.*, p<.05; **, p <0.01. ^#^, Data of one patient is missing; ^¶^, Data of two patients is missing.

### Concentration of ketamine and its metabolites in plasma

The highest median ketamine (median: 34.80 ng/ml; min: 12.9, max: 162.4 ng/ml) plasma concentration was observed 30 minutes after administration. Norketamine was the major metabolite during all timepoints at the day of administration with norketamine concentrations exceeding ketamine concentrations as early as 20 minutes after administration. Median norketamine plasma concentration was highest (median: 290.70 ng/ml; min: 174.7, max: 483.7 ng/ml) 60 minutes after administration. Ketamine concentration was below the quantification threshold in all samples obtained after 24 hours and measured 48 hours after the application of the fifth dose (day 11). Norketamine concentration after 24 hours and 11 days after drug administration was below the quantification threshold in more than 50% of subjects. Dehydronorketamine demonstrated significant instability in blood samples stored at −80°C rendering absolute results unreliable. Highest median dehydronorketamine concentration was measured at 120 minutes (median: 58.40; min: 19.1, max: 254.1 ng/ml). Information on missing samples and results of exploratory analyses including dehydronorketamine are reported in the supplement.

### Exploratory analyses

A correlation between the CADSS total score obtained 30 minutes after the first drug administration and MADRS change after seven days (r_s_ = −0.61, p < 0.01) was observed (see Supplemental Figure 1).Furthermore, a correlation between the CADSS total score and plasma levels of ketamine (R = 0.63, p = 0.0017) and norketamine (R = 0.42, p = 0.049) after 30 minutes and ketamine after 60 minutes (R = 0.58, p = 0.0056) was observed. Further exploratory correlation analyses are reported in the supplement.

### Meta-analysis of RCTs assessing antidepressant efficacy of oral ketamine

The systematic literature research yielded seven eligible RCTs (Arabzadeh et al., 2018; Colla et al., 2024; Domany et al., 2019; Jafarinia et al., 2016; Seraj et al., 2025; Smith-Apeldoorn et al., 2024). Together with the present investigation the meta-analysis comprised eight studies with a total of 592 patients. The summary effect estimate for differences in depression rating scales between groups after treatment clearly indicated antidepressant efficacy of oral ketamine treatment (standardized mean difference = −0.80, 95% CI: [−1.43, −0.17]). Furthermore, efficacy was evident with regard to differences in the number of responders between treatment groups. The NNT estimated by meta-analysis was 4.89 (95% CI: [3.41, 8.66]) for response and 9.10 (95% CI: [5.43, 28.03] for remission (Figure 2).

**Figure 2.**
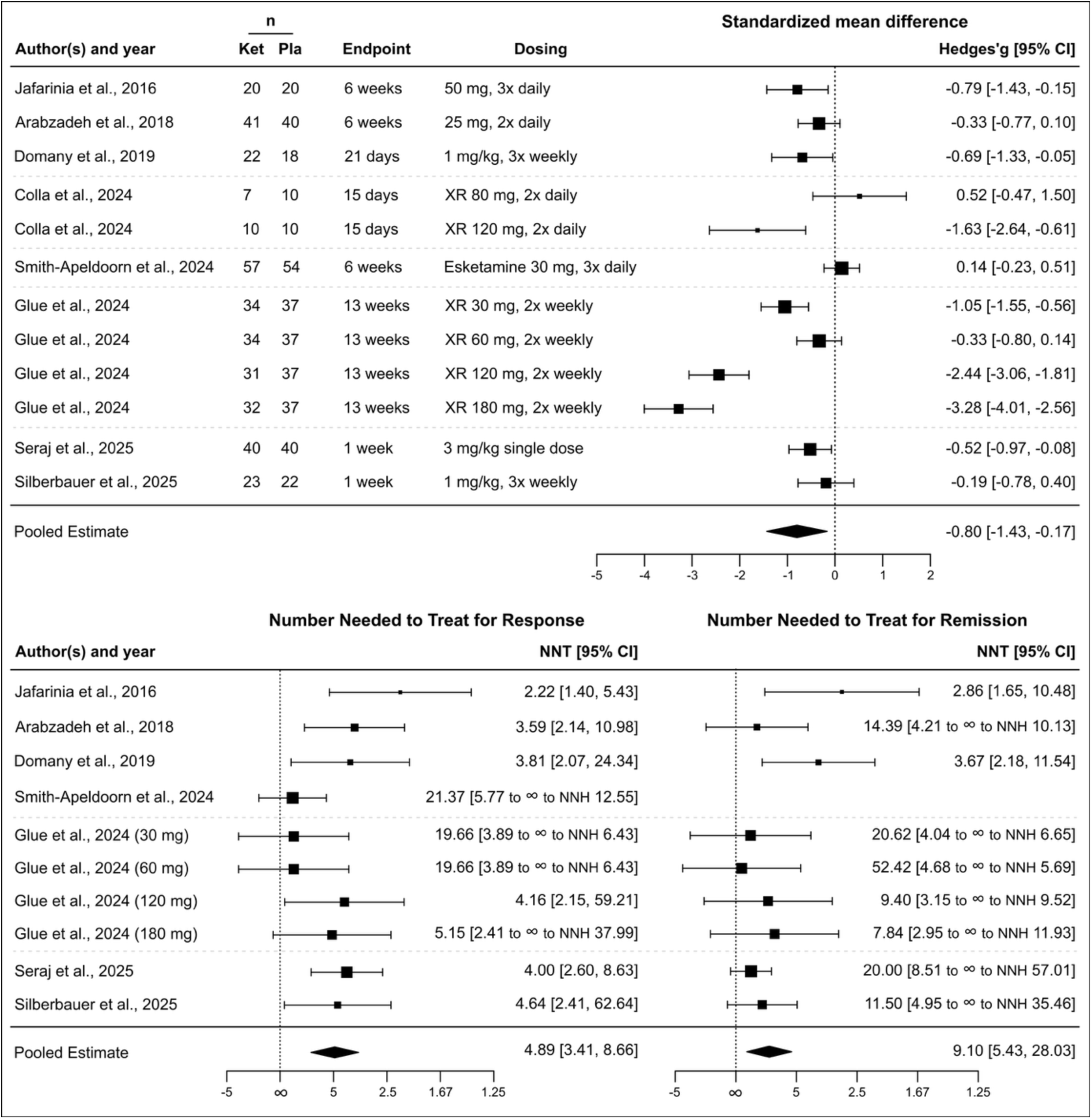
Antidepressant efficacy of peroral ketamine treatment in depression. Top panel: A forest plot summarizing the differences in depression rating scores between groups receiving ketamine versus control treatments, yielding an effect size of −0.80 (95% CI: [−1.43, −0.17]). Bottom panel: Forest plots illustrating the number needed to treat (NNT) for response (left) and remission (right). The NNT for response is 4.89 (95% CI: [3.41, 8.66]), while the NNT for remission is 9.10 (95% CI: [5.43, 28.03]).

## DISCUSSION

### Main findings

This randomized controlled trial assessed the efficacy and tolerability of repeated oral ketamine in patients suffering from depression. While the primary endpoint of difference in MADRS scores between treatment groups after 7 days was not significant, the difference in response rates is considered as clinically relevant. Oral ketamine was well tolerated and occurring side effects were mild and transient.

Our meta-analysis of randomized controlled trials comprising a total of 592 patients demonstrated antidepressant efficacy of oral ketamine treatment.

### Antidepressant efficacy

Evidence on antidepressant efficacy of oral ketamine is currently limited and, so far, only one randomized controlled study demonstrated antidepressant efficacy of repeated oral ketamine in a study design that directly compares to the present investigation (Domany et al., 2019). The reported mean reduction of ∼10 points on the MADRS score after 14 days is consistent with our results (mean reduction after 11 days: 9.95). Differences in observation periods and a lower response rate in the control group in the study by *Domany et al.* may explain variations in comparison to our study. In the trial by *Jafarinia et al.*, oral ketamine was compared to the analgesic diclofenac in depressed patients with chronic pain (Jafarinia et al., 2016). A statistically significant effect was reported after six weeks, which exceeds our observational period and may be confounded by analgesic effects of ketamine. Add-on treatment of daily oral ketamine to a newly initiated selective serotonin reuptake inhibitor accelerated treatment response in the RCT by *Arabzadeh and colleagues (Arabzadeh et al., 2018)*. Patients in our study, in contrast, received stable psychopharmacological treatment for two weeks and ketamine was administered less frequently. Our findings align with recent data from RCTs showing a positive but non-significant trend for antidepressant efficacy of low-dose peroral ketamine (Colla et al., 2024; Smith-Apeldoorn et al., 2024). Both trials suggest that higher doses or optimized dosing strategies may enhance clinical outcomes. Promising results were recently reported by *Glue et al.* using an extended-release oral ketamine formulation in a TRD population (Glue et al., 2024). Their use of an enrichment design successfully addressed the challenge to separate clinical response between groups as observed in our trial. Maintaining the blind is a key challenge in ketamine trials give its unambiguous subjective effects. Midazolam, frequently used as a control condition in studies demonstrating antidepressant efficacy of intravenous ketamine, offers comparable pharmacokinetics and behavioral effects (Murrough et al., 2013; Phillips et al., 2019). So far, only one randomized controlled trial investigating oral ketamine has employed an active control condition, i.e. midazolam (Seraj et al., 2025). In this study, *Seraj et al.* demonstrated a rapid reduction in suicidal ideation following a single dose of oral ketamine in a carefully selected sample of MDD patients without comorbidities. Despite the use of a threefold higher dose (3mg/kg), the observed antidepressant effects were modest, with a response rate of approximately 25% at one-week follow-up—a finding that closely aligns with the results of our study. However, increases in response rates observed after 11 days of repeated ketamine administration in our trial could not be captured by *Seraj et al.*, given their single-dose design and shorter observation period.

The placebo effect in depression trials is known to be large (Jones et al., 2021) and greater improvement in depression severity following midazolam vs. saline and resulting smaller effect sizes for ketamine were shown (Wilkinson et al., 2019). Thus, the use of midazolam as an active control may have prevented our results from reaching statistical significance. However, the observed mean reduction in MADRS (from 6.07 after 24 hours to 9.95 after 11 days) aligns with previously reported treatment effects in positive ketamine trials using intravenous and oral administration, respectively, and can be considered as clinically meaningful (Domany et al., 2019; Phillips et al., 2019; Turkoz et al., 2021). However, our findings are also in line with recent reports of low effect sizes observed in studies successfully masking treatment allocation through surgical anesthesia (Lii et al., 2023). Clinical baseline characteristics, such as high anxiety levels (12 patients with anxious depression in the midazolam group vs. 10 in the ketamine group according to the HDRS anxiety-somatization factor), and patients’ expectancy may have contributed to the large response in the midazolam group. In sum, the study designs of previous RCTs were highly heterogeneous and, consequently, we conducted a meta-analysis incorporating previously published RCTs in order to contextualize our results within this diverse data landscape. Our results strongly support antidepressant efficacy of oral ketamine treatment in terms of symptom reduction as well as differences in response and remission rates between treatment groups. Consequently, we argue that oral ketamine treatment may represent a viable option for the treatment of depression.

### Tolerability

Previous studies reported elevation in dissociative measures in the treatment of depression using sub-anesthetic ketamine (Fava et al., 2018; Murrough et al., 2013). In our study, statistically significant higher dissociation scores on the CADSS were observed in the ketamine group. However, this increase was modest compared to previous studies using intravenous ketamine (Fava et al., 2018; Murrough et al., 2013) and comparable to those reported with intranasal esketamine (Chen et al., 2022; Fedgchin et al., 2019). Evidence on whether dissociation correlates with the immediate or long-lasting antidepressant effects of antidepressants remains inconclusive (McIntyre et al., 2021). In our study, we observed a significant correlation between MADRS reduction after 7 days and dissociation after the first drug administration as measured using the CADSS. We observed significant group differences in the domain vigilance reduction and the subscale disembodiment of the ASC between groups. In line with our results, ketamine induced dissociation is typically captured by the subscale disembodiment (Marguilho et al., 2023). With the majority of subscales ranging below the 10 per cent mark of the scalés maximum, oral ketamine induced lower scores on the ASC when compared to intravenous drug administration (Passie et al., 2021; Vidal et al., 2018) which suggests good tolerability of this formulation. In contrast to previous trials, side effects were assessed quantitatively in a structured manner using an extensive visual analogue scale (Domany et al., 2019). All observed side effects were mild and transient. Most commonly, dry mouth, dizziness, poor coordination, poor concentration and fatigue were reported (see Table 2). Importantly, no cumulative effect was observed after repeated dosing. A significant difference after repeated dosing was only observed for one item (poor coordination) suggesting habituation to drug effects. We did not observe lower urinary tract symptoms in our trial. However, given the dose-response relationship between ketamine exposure and these symptoms (Habert et al., 2016), this potential adverse event should be considered during long-term treatment with oral ketamine, which may require higher doses when compared to intravenous or intranasal formulations.

### Dosing and pharmacokinetics

We used a fixed dose of 1mg/kg bodyweight, a dosing regimen that yielded promising results in the trial by *Domany et al.* (Domany et al., 2019). The low bioavailability is frequently cited as an impediment to oral ketamine treatment and, indeed, ketamine plasma levels observed in our study were lower than the levels reported in studies following intravenous administration (Zarate et al., 2012a). Peroral administration of ketamine leads to pronounced first-pass metabolism, resulting in norketamine as the primary circulating compound at steady state (Glue et al., 2021). Formulations with extensive metabolism and lower peak ketamine plasma concentrations including peroral ketamine were demonstrated to be safer and were associated with better tolerability (Dutton et al., 2023; Glue et al., 2021). Results from the study by *Glue et al.* suggest a dose-response relationship, with significant antidepressant effects observed at doses of 120 mg during the open-label phase and 180 mg in the double-blind phase (Glue et al., 2024). While these doses exceed those used in our study by factors of 1.7 and 2.6 for a person with 70kg bodyweight, respectively, the findings also suggest that relatively low doses of approximately 2 mg/kg may yield a meaningful antidepressant effect. However, a head-to-head comparison of 1 mg/kg and 2 mg/kg found no significant advantage of the higher dose in terms of efficacy, but observed a higher incidence of adverse effects, thereby supporting the use of the lower dose for safety reasons (KheirAbadi and Golkar, 2023).

### Implications of peroral ketamine treatment

Peroral ketamine treatment for depression offers promising benefits. Unlike intravenous administration, oral administration simplifies clinical procedures, eliminating the need for intravenous cannulation and, thus, reducing patient time in clinics. While a single intranasal dose of esketamine costs over €550, a full treatment series using oral ketamine in our trial may total less than €10 for an average 70 kg person, making it a highly affordable alternative. However, the potential for abuse necessitates cautious implementation of at-home administration with clear guidelines from healthcare providers and regulators. If carefully managed, peroral ketamine could greatly broaden access to effective depression care within communities.

### Strengths and Limitations

This is the first study to assess antidepressant efficacy of repeated oral ketamine using midazolam as an active comparator (Meshkat et al., 2023). We conducted a comprehensive assessment of antidepressant efficacy and safety. Side effects were systematically evaluated after single and repeated dosing using clinician-rating and self-rating questionnaires. Additionally, we present pharmacokinetic data obtained from serial assessments of plasma levels of ketamine and its metabolites. In- and exclusion criteria were chosen to ensure enrollment of a clinically representative population. Subjects with uni- and bipolar depression were enrolled and only minor restrictions with respect to psychiatric and medical comorbidities were made. The heterogenous study population, however, complicates the comparability of our results with studies comprising a strictly selected patient population, such as patients with TRD. Unfortunately, we did not measure patients’ treatment expectancy prior to randomization. Moreover, the relatively short observational period and the lack of long-term follow-up data may be considered as a limitation of our study.

### Conclusion

In conclusion, our findings, supported by meta-analytic evidence, provide strong support for antidepressant efficacy and safety of peroral ketamine and highlight the need for future research on optimal dosing and delivery methods.

## DATA AVAILABILITY STATEMENT

Raw data will not be publicly available due to reasons of data protection. Processed data can be obtained from the corresponding author with a data-sharing agreement, approved by the departments of legal affairs and data clearing of the Medical University of Vienna.

## CRediT AUTHORSHIP CONTRIBUTION STATEMENT

**Leo R. Silberbauer:** Conceptualization, Data curation, Formal analysis, Funding acquisition, Investigation, Methodology, Project administration, Validation, Visualization, Writing – original draft, Writing – review and editing. **Benjamin Eggerstorfer:** Formal analysis, Investigation, Project administration, Visualization, Writing – review and editing. **Paul Michenthaler:** Investigation, Writing – review and editing. **Sabine Reichel:** Formal analysis, Resources, Writing – review and editing. **Thomas Stimpfl:** Formal analysis, Resources, Supervision, Writing – review and editing. **Thomas Vanicek:** Investigation, Supervision, Writing – review and editing. **Angela Naderi-Heiden:** Investigation, Supervision, Writing – review and editing. **Siegfried Kasper:** Investigation, Resources, Supervision, Writing – review and editing. **Rupert Lanzenberger:** Project administration, Resources, Supervision, Writing – review and editing. **Gregor Gryglewski:** Conceptualization, Data curation, Formal analysis, Funding acquisition, Investigation, Methodology, Project administration, Supervision, Validation, Writing – original draft, Writing – review and editing.

## COMPETING INTEREST

RL received travel grants and/or conference speaker honoraria from Bruker BioSpin within the last three years and investigator-initiated research funding from Siemens Healthcare regarding clinical research using PET/MR. He is a shareholder of the start-up company BM Health GmbH since 2019. SK received grants/ research support, consulting fees and/or honoraria within the last three years. He received grant/ research support from Lundbeck; he has served as a consultant or on advisory boards for Celegne, IQVIA, Janssen, Lundbeck, Mundipharma, Recordati, Takeda and Schwabe; and he has served on speaker bureaus for Angelini, Aspen Farmaceutica S.A., Janssen, Krka Pharma, Lundbeck, Medichem Pharmaceuticals Inc., Neurax-pharma, OM Pharma, Pierre Fabre, Sanofi, Servier, Schwabe, Sun Pharma. The remaining authors declare no potential conflict of interest with respect to the research, authorship, and/ or publication of this article.

## ACKNOWLEDGEMENTS

We thank Patricia Anna Handschuh, Alim Emre Başaran, Mathis Godber Godbersen, Jakob Felix Unterholzner, Valentin Popper, Marius Hienert and Richard Frey for their medical support. This research was funded in part by the Medical-Scientific Fund of the Mayor of the City of Vienna. LRS and GG were recipients of DOC fellowships of the Austrian Academy of Sciences at the Department of Psychiatry and Psychotherapy, Medical University of Vienna.

## Notes

### Clinical Trial

NCT02992496

### Author Declarations

All study related procedures were reviewed and approved by the ethics committee of the Medical University of Vienna and carried out in accordance with ethical principles that have their origin in the Declaration of Helsinki.

### Summary of Updates

Minor textual adjustments to reduce the length of the manuscript.

