## Supplemental Material for "Oral Ketamine for the Treatment of Depression: A randomized controlled trial and meta-analysis"

### *Supplemental information*

### *Contents*

**Supplemental Text.** Supplementary methods……………………………………………………………………..2
**Supplemental Table S1.** Response and remission rates…………...…………………………………………...3
**Supplemental Table S2.** Results of the CADSS and the ASC scale……………………………………………..4
**Supplemental Table S3.** Plasma levels of ketamine and metabolites……………………………...……..5
**Supplemental Text.** Supplementary results….…………………………………………..…………….…….6
**Supplemental Figure S1.** Association between MADRS change and dhnk plasma levels/CADSS score.………7
**Supplemental Figure S3.** Consort Flow Diagram….………………………………………..………………8
**Supplemental Figure S4.** PRISMA flow diagram……………………………….……………………………....9
**Supplemental References**………………………………..……………………………………………………10

**Supplementary methods**

Sample size

Power calculation was performed using G*Power 3.1.9.2. An effect size of Cohen's d=0.5 and targeted power of 95% resulted in a required total sample size of 55 patients. However, the study was discontinued during the coronavirus pandemic before reaching the intended sample size.

Randomization

Subjects were randomized using block randomization with variable-sized blocks of 6 to 16 subjects using R (https:// www.R-project.org/). Within each block, subjects were evenly assigned to receive either ketamine or midazolam. Randomization was performed by an independent researcher of our group not involved in study conduction and analysis.

Study medication

The blinded study medication was diluted using 5% glucose solution and provided by the hospital pharmacy using syringes containing 10mL solution. Ketamine was provided at a concentration of 10mg/ml and midazolam at a concentration of 0.3mg/ml. One mL per 10kg bodyweight from the prepared solution was transferred to a cup by the study physician. Participants were instructed to ingest the solution at once and were provided with a cup of water after ingestion. After administration of the medication, participants were continuously monitored by the study physician at the Department of Psychiatry and Psychotherapy at the Medical University of Vienna.

Blood sampling

To investigate the association between antidepressant response and tolerability, respectively, with plasma levels of ketamine and its metabolites, serial venous blood samples were drawn from the cubital vein at 20, 30, 60, 120 minutes and 24 hours after administration of the first drug dose. Moreover, a sample was obtained after the last dose of the study medication.

Meta-analysis of RCTs assessing antidepressant efficacy of oral ketamine

The following search terms were used in various combinations: ("oral ketamine" OR "peroral ketamine") AND ("depression" OR "MDD" OR "major depressive disorder" OR "major depressive episode"). In PubMed, search terms were applied to the title and abstract fields ([tiab]) and supplemented with MeSH terms for "depression." In Scopus, the search was conducted within the Title, Abstract, and Keywords fields. In PsycINFO (via Ovid), a broad field search was used, targeting multiple relevant fields (.mp.), which include titles, abstracts, subject headings, and keywords. In Web of Science, the search was performed in the Topic field (TS=), which encompasses titles, abstracts, author keywords, and Keywords Plus®. Studies were included if they fulfilled the following inclusion criteria: peer-reviewed English or German language original articles, randomized controlled trials, studies that reported means and standard deviation values or effect sizes of antidepressant outcome measures. Exclusion criteria were case reports, reviews, meta-analyses and preclinical studies. The literature search was conducted in accordance with the guidelines of the PRISMA statement (Page et al., 2021), and the selection of included studies was performed using the Rayyan software (Ouzzani et al., 2016). We included the results of the present trial. Analyses were performed using the metafor package (Viechtbauer, 2010). This meta-analysis was not preregistered, as it was performed post hoc to provide contextual interpretation of the data collected within this clinical trial.

| | | | | |
|---|---|---|---|---|
| **<u>24 hours</u>** | | | | |
| **Response** | | | | |
| | Non response | Response | n | p |
| Midazolam | 22 (100%) | 0 (0%) | 22 | |
| Ketamine | 22 (95.65%) | 1 (4.35%) | 23 | 1 |
| **Remission** | | | | |
| | Non remission | Remission | n | |
| Midazolam | 22 (100%) | 0 (0%) | 22 | |
| Ketamine | 22 (95.65%) | 1 (4.35%) | 23 | 1 |
| **<u>7 days</u>** | | | | |
| **Response** | | | | |
| | Non response | Response | n | |
| Midazolam | 21 (95.46%) | 1 (4.55%) | 22 | |
| Ketamine | 17 (73.91%) | 6 (26.10%) | 23 | 0.10 |
| **Remission** | | | | |
| | Non remission | Remission | n | |
| Midazolam | 22 (100%) | 0 (0%) | 22 | |
| Ketamine | 21 (91.30%) | 2 (8.70%) | 23 | 0.49 |
| **<u>11 days</u>** | | | | |
| **Response** | | | | |
| | Non response | Response | n | |
| Midazolam | 20 (90.91%) | 2 (9.10%) | 22 | |
| Ketamine | 15 (65.22%) | 8 (34.78%) | 23 | 0.07 |
| **Remssion** | | | | |
| | Non remission | Remitter | n | |
| Midazolam | 21 (95.46%) | 1 (4.55%) | 22 | |
| Ketamine | 18 (91.30%) | 2 (8.70%) | 23 | 1 |

**Table S1** Response and remission rates are given for 24 hours, 7 and 11 days after the first drug administration. Total number of patients with available data at each timepoint (n) and p-values from Fisher´s exact test are reported.

| | **Midazolam** (mean±sd) | **Ketamine** (mean±sd) | t | p |
|---|---|---|---|---|
| CADSS | n=22 | n=23 | | |
| Total score | 2.36±4.22 | 8.65±9.95** | -2.78 | <0.01** |
| 5 Dimensions Altered States of Consciousness (5D-ASC) scale | n=19 | n=21 | | |
| Oceanic boundlessness | 6.86±9.84 | 12.15±15.94 | -1.28 | 0.21 |
| Anxious ego dissolution | 6.10±12.30 | 8.21±8.24 | -0.63 | 0.53 |
| Visionary restructuralization | 4.99±8.73 | 8.42±12.14 | -1.03 | 0.31 |
| Auditory alterations | 2.69±4.09 | 5.81±10.35 | -1.27 | 0.21 |
| Vigilance Reduction | 15.55±17.34 | 31.44±22.56* | -2.51 | <0.05* |
| Experience of Unity | 5.66±10.14 | 9.44±17.49 | -0.84 | 0.41 |
| Spiritual Experience | 4.23±10.44 | 7.05±12.92 | -0.76 | 0.45 |
| Blissful State | 10.21±13.00 | 15.86±20.88 | -1.04 | 0.31 |
| Insightfulness | 6.28±13.42 | 7.05±13.79 | -0.18 | 0.86 |
| Disembodiment | 4.33±8.22 | 17.32±23.74* | -2.36 | <0.05* |
| Impaired Control and Cognition | 7.44±12.01 | 10.21±11.86 | -0.73 | 0.47 |
| Anxiety | 4.52±11.08 | 6.10±5.91 | -0.55 | 0.58 |
| Complex Imagery | 4.97±8.43 | 8.75±14.84 | -1.00 | 0.32 |
| Elementary Imagery | 3.30±7.45 | 7.64±16.23 | -1.10 | 0.28 |
| Audio-Visual Synesthesia | 2.53±3.77 | 8.41±16.23 | -1.62 | 0.12 |
| Changed Meaning of Percepts | 7.70±14.50 | 6.05±13.14 | 0.38 | 0.71 |

**Table S2** Results of the Clinician Administered Dissociative States Scale (CADSS) and the Altered States of Consciousness (ASC) scale. Results of the 5D-ASC are shown in percentage of scale maximum. n, number of subjects; sd, standard deviation; *, p<.05; **, p <0.01.

| **Sampling time** | **Ketamine** Median (MAD) | | **Norketamine** Median (MAD) | | **Dehydronorketamine** Median (MAD) | |
|---|---|---|---|---|---|---|
| 20 min | 31.65 (23.94) | 22/22 | 216.55 (197.85) | 22/22 | 15.95 (16.31) | 22/22 |
| 30 min | 34.80 (16.38) | 22/22 | 273.0 (134.47) | 22/22 | 37.30 (36.32) | 22/22 |
| 60 min | 29.20 (16.31) | 21/22 (1 NA) | 290.70 (57.97) | 21/22 (1 NA) | 49.40 (40.48) | 21/22 (1 NA) |
| 120 min | 19.90 (12.45) | 20/22 (2 NA) | 213.65 (64.94) | 20/22 (2 NA) | 58.40 (36.92) | 20/22 (2 NA) |
| 24h | Not detected | 19/22 (20 n.d., 2 NA) | 0 (0) | 9/22 (11 n.d., 2 NA) | 4 (2.15) | 18/22 (2 n.d., 2 NA) |
| Day 11 | Not detected | 20/22 (20 n.d., 2 NA) | 0 (0) | 2/22 (18 n.d., 2 NA | 0 (0) | 7/22 (13 n.d., 2 NA) |

**Table S3** displays plasma concentration of ketamine, norketamine and dehydronorketamine. Assessment of plasma levels of ketamine and its metabolites was performed in 22 out of 23 subjects in the ketamine group. Missing plasma samples (NA) and values below the quantification threshold (n.d.) are reported. Dehydronorketamine demonstrated significant instability in blood samples stored at -80 degrees Celsius, rendering absolute results unreliable.

**Supplementary results**

Exploratory analyses

Correlation between MADRS reduction and plasma levels

No correlation between ketamine and norketamine plasma levels and MADRS reduction was observed. The association between MADRS reduction and dehydronorketamine plasma levels is shown in Figure S1.

Correlation between MADRS reduction and the ASC

A significant correlation between MADRS reduction after seven days and the dimensions visionary restructuralization (R = -0.65, p = 0.0027) and auditory alterations (R = -0.64, p = 0.003), and the subdimensions experience of unity (R= -0.46, p = 0.045), spiritual experience (R = -0.62, p = 0.005), blissful state (R = -0.67, p = 0.0018), insightfulness (R = -0.53, p = 0.021), complex imagery (R = -0.63, p = 0.004), elementary imagery (R = -0.52, p = 0.021) and audio-visual synesthesia (R = - 0.5, p = 0.03), respectively, was observed.

Correlation between ASC and plasma levels

A correlation between ketamine plasma concentration 20 minutes after administration and the dimension oceanic boundlessness (R = 0.5, p = 0.02), the subdimension anxiety (R = 0.51, p = 0.018) and elementary imagery (R = 0.57, p = 0.0069) and, respectively, ketamine plasma concentration 30 minutes after administration and the dimensions oceanic boundlessness (R = 0.61, p = 0.0037), anxious ego-dissolution (R = 0.49, p = 0.024), visionary restructuralization (R = 0.49, p = 0.025), the 5D-ASC total score (R = 0.5, p = 0.023) and the 3D-OAV total score (R = 0.49, p = 0.026), the subdimension disembodiment (R = 0.5, p = 0.021), anxiety (R = 0.56, p = 0.0089) and elementary imagery (R = 0.46, p = 0.035) was observed.

Norketamine plasma concentration after 20 minutes correlated with the subdimension elementary imagery (R = 0.46, p = 0.037) and norketamine plasma levels after 30 minutes correlated with the subdimensions disembodiment (R = 0.45, p = 0.039) and anxiety (R = 0.47, p = 0.033).

A correlation between dehydronorketamine plasma concentration 30 minutes after administration and the subdimension changes meaning of percepts (R = -0.44, p = 0.044); dehydronorketamine plasma concentration 60 minutes after administration and the dimension anxious ego-dissolution (R = -0.45, p = 0.044); dehydronorketamine plasma concentration 120 minutes after administration and the dimension anxious ego-dissolution (R = -0.49, p = 0.035), visionary restructuralization (R = -0.49, p = 0.034) and auditory alterations (R = -0.47, p = 0.041) was observed.

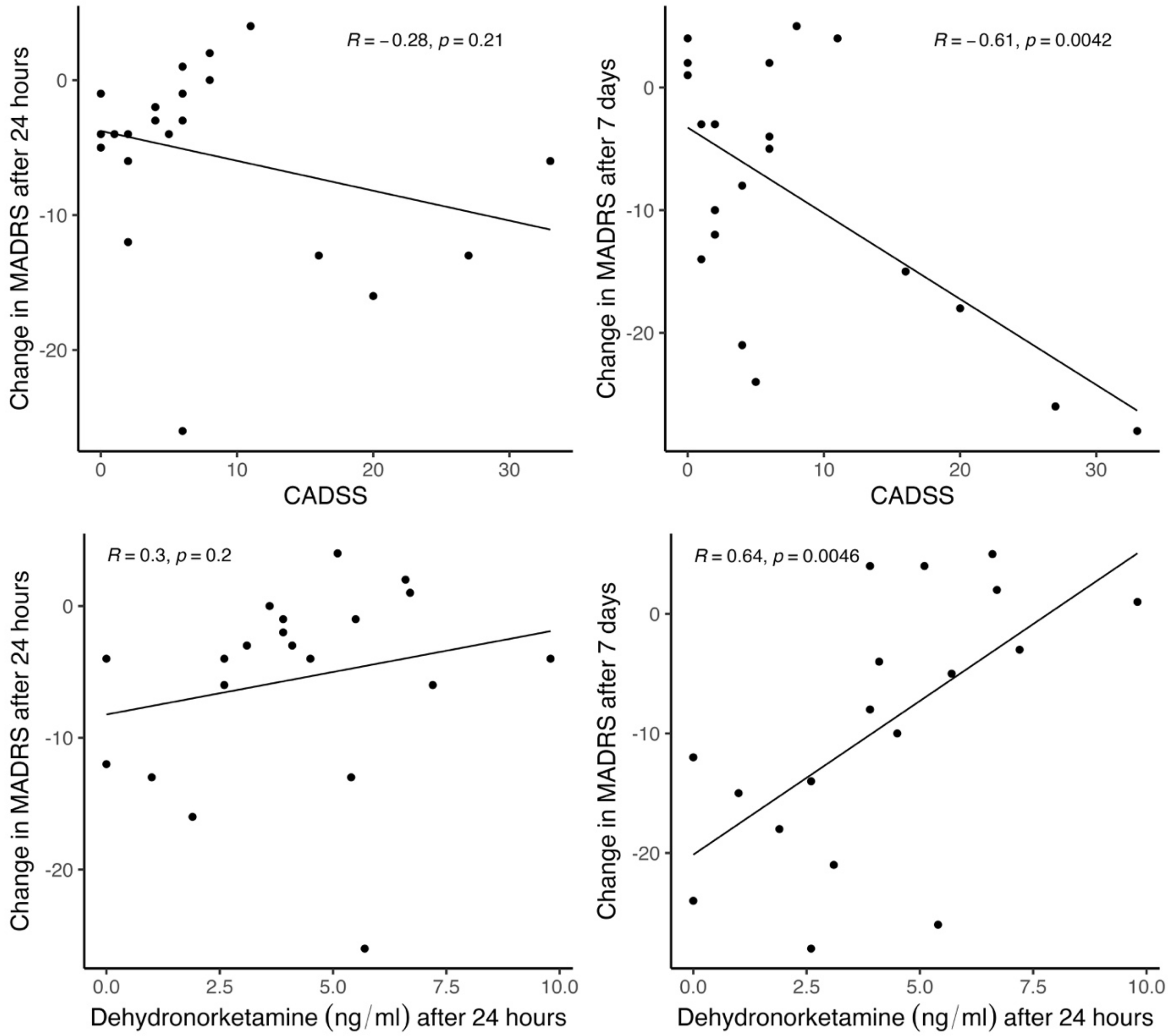

**Figure S1.** Top row: The association between the Clinician Administered Dissociative States Scale (CADSS) score and change in MADRS after 24 hours and 7 days (primary endpoint) in the ketamine group is shown. Bottom row: Scatterplots display the association between mean change of MADRS total score after 24 hours and 7 days (primary endpoint), respectively, and dehydronorketamine plasma levels measured 24 hours after the first drug administration.

CONSORT
TRANSPARENT REPORTING of TRIALS

**CONSORT 2010 Flow Diagram**

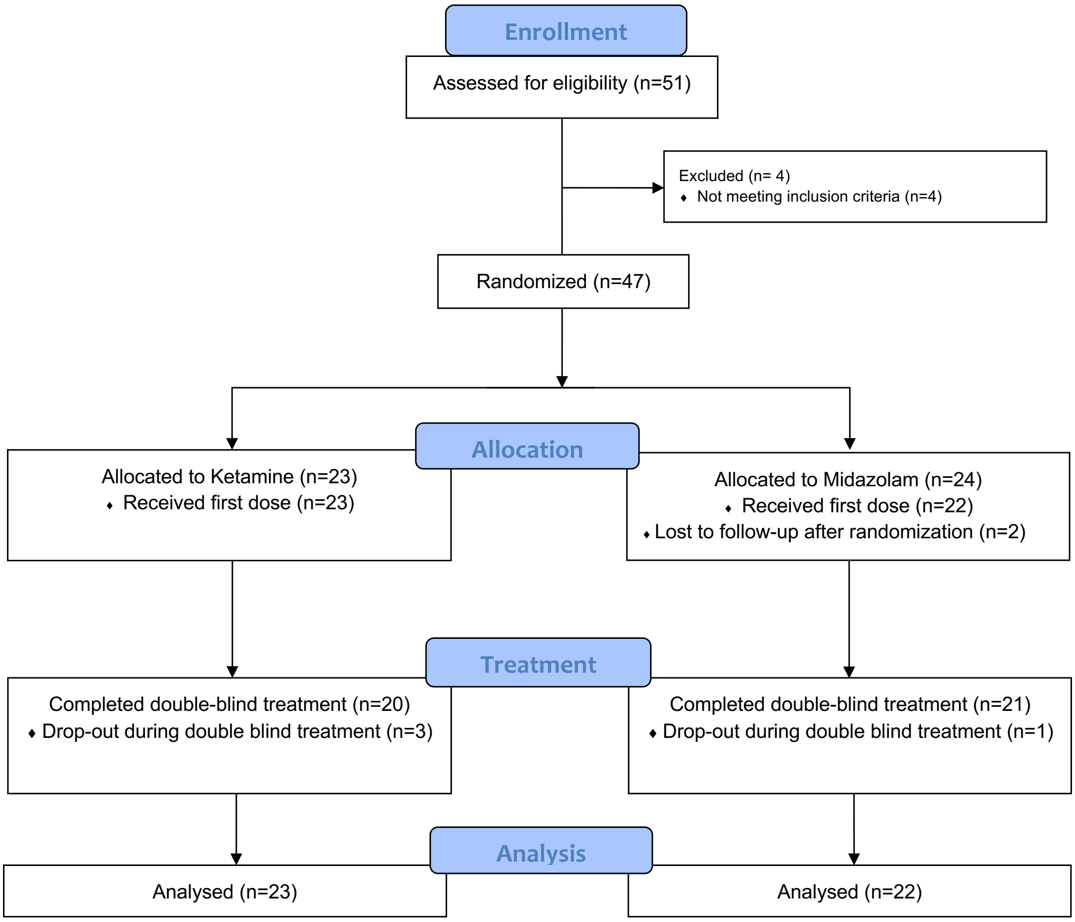

**Figure S3** Consort flow diagram

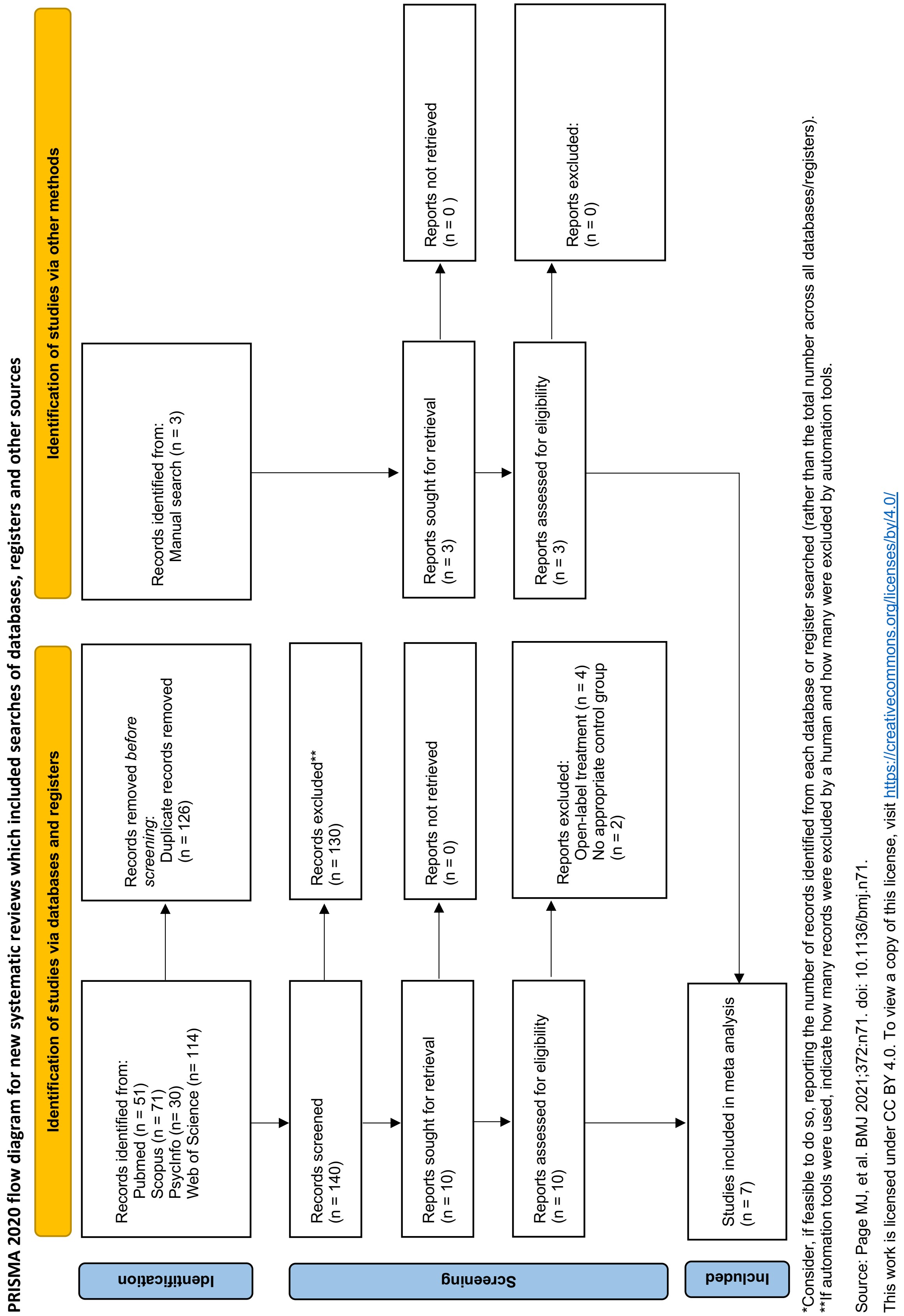

**Figure S4** PRISMA flow diagram
